# Methodological review to develop a list of bias items for adaptive clinical trials: Protocol and rationale

**DOI:** 10.1101/2024.04.25.24306353

**Authors:** Phillip Staibano, Tyler McKechnie, Alex Thabane, Daniel Olteanu, Keean Nanji, Han Zhang, Carole Lunny, Michael Au, Michael K. Gupta, Jesse D. Pasternak, Sameer Parpia, JEM (Ted) Young, Mohit Bhandari

## Abstract

**Background:** Randomized-clinical trials (RCTs) are the gold-standard for comparing health care interventions, but can be limited by early termination, feasibility issues, and prolonged time to trial reporting. In contrast, adaptive clinical trials (ACTs), which are defined by pre-planned modifications and analyses that occur after starting patient recruitment, are gaining popularity as they can streamline trial design and time to reporting. As adaptive methodologies continue to be adopted by researchers, it will be critical to develop a risk-of-bias tool that evaluates the unique methodological features of ACTs so that their quality can be improved and standardized for the future. In our proposed methodological review, we plan to develop a list of risk-of-bias items for ACTs to develop a candidate instrument.

**Methods and analysis:** We will perform a systematic database search to capture: (1) ACTs published in any discipline of medicine and/or surgery; and (2) studies that have proposed or reviewed items pertaining to methodological risk, bias, and/or quality in ACTs. We will perform a comprehensive search of citation databases, such as Ovid MEDLINE, EMBASE, CENTRAL, the Cochrane library, and Web of Science, in addition to multiple grey literature sources to capture published and unpublished literature related to ACTs and studies evaluating the methodological quality of ACTs. We will also search methodological registries for any risk of bias tools for ACTs. All screening and review stages will be performed in duplicate with a third senior author serving as arbitrator for any discrepancies. Included ACTs will be analyzed in a descriptive manner, and we will perform regression analysis to identify factors associated with poor reporting quality and high risk of bias. We will also perform a risk of bias assessment of ACTs using the Cochrane risk-of-bias 2.0 tool and we will assess reporting quality using the CONSORT-ACE tool. These assessments will be performed independently and in duplicate. This will be done to help generate risk of bias concepts, themes, and items that can be included in the candidate tool. For all studies of methodological quality and risk of bias, we will extract all pertinent bias items and/or tools. We will combine conceptually similar items in a descriptive manner and classify them as referring to bias or to other aspects of methodological quality, such as reporting. We will plan to generate pertinent risk of bias items and fields and finally, a candidate tool that will undergo further refinement, testing, and validation in future development stages.

**Ethics and dissemination:** This review does not require ethics approval as human subjects are not involved. As mentioned previously, this study is the first step in developing a tool to evaluate the risk of bias and methodological quality of ACTs.

## INTRODUCTION

Evidence-based medicine has revolutionized the development of clinical practice guidelines and decision making in healthcare [1]. Randomized-controlled trials (RCTs) are the gold-standard for comparing the effectiveness and safety of novel healthcare interventions [2]. Conventional RCTs, however, can be burdened by high costs, early termination due to feasibility issues, and an overly rigid design that does not permit adjustments for unforeseen challenges [3]. These issues are amplified in surgical trials and as such, the annual number of published surgical trials remains stagnant [4, 5]. As a response to these challenges, researchers have begun using adaptive trial designs, which allow for dynamic protocol changes after beginning patient recruitment. Adaptive clinical trials (ACTs) Adaptive clinical trial (ACTs) are a novel approach may help to streamline patient recruitment, drop poorly performing treatment arms, and combine clinical trial stages, thereby, reducing unnecessary trial costs, decreasing sample size, and accelerating time to reporting [6]. For instance, the TAILoR trial of telmisartan in HIV employed an interim analysis at half maximal patient recruitment and dropped the most ineffective medication dosage group based on a pre-specified efficacy threshold [7]. Adaptive methodologies helped to streamline the clinical trial process during COVID-19 to optimize the number of therapies evaluated and reduce uncertainties related to the natural history of the disease and the number of patients necessary for each trial [8]. Adopting ACTs when prolonged RCTs are impractical may also reduce needed funding, thereby overcoming barriers to conducting trials in developing nations [9]. Stakeholders, however, report that adaptive trial designs remain nebulous with practical barriers, including high bias potential, ethical concerns, and a lack of knowledge dissemination amongst trialists [10].

In 2020, CONSORT published an extension for adaptive trials to guide ACT reporting [11]. These guidelines ACT-specific methodological components such as pre-planned interim analyses and sample size estimation (and re-estimation) descriptions [11]. In conjunction with CONSORT reporting guidelines, a validated risk-of-bias tool developed in a similar manner to the Cochrane risk-of-bias 2.0 tool [12], may improve the design of ACTs and the quality of future meta-analyses combing ACTs. Risk-of-bias tools are designed for specific study designs (e.g. RCTs) and help to promote methodological transparency and reproducibility while minimizing bias, so results can be accurately interpreted and soundly applied to patient care. For conventional RCTs, there exist several tools and checklists to guide reporting or evaluate quality and bias risk (Table 1) [12-25]. There is no existing risk-of-bias tool to evaluate the methodological limitations of ACTs, which is of particular importance due to the potential for ACTs to be impacted by bias if not soundly designed [26]. It is for this reason that we have decided to embark upon creating a novel risk-of-bias tool to improve the quality of future ACTs and ACT meta-analyses.

**Table 1.** Tools and checklists to aid in RCT conduct, or to assess the reporting or methodological quality of an RCT. Adapted from Lunny et al. (2021).

| **Tool purpose** | **Examples of tools or checklists** | **Description of the example tool or checklist** | **Available tool for ACTs** |
| --- | --- | --- | --- |
| **Assess the quality of published RCTs** | CASP-RCT [22], CEBM-RCT [14], Jadad scale [13], JBI checklist [23], LEGEND [25], TRACT [15], SIGN [24] | These tools to assist in conducting and reviewing published RCTs that consist of multiple domains addressing trial design and internal validity, result reporting, and interpretability. | None |
| **Assess the risk of bias of published RCTs** | Cochrane RoB tool (2.0) [11] | Cochrane RoB 2.0 is a five-domain domain tool that evaluates bias arising from randomization, protocol deviations, missing outcome data, outcome measurement, and result reporting. Studies are given an overall bias risk score of low, high, or some concerns. | None |
| **Assess the external validity of RCTs** | None [18] | None | None |
| **Guidance for the complete reporting of RCTs** | CONSORT [19] | CONSORT is a 25-item checklist that assists with transparent reporting of RCT design, analysis, and interpretation. | CONSORT-ACE [10] |
| **Guidance checklist for clinical readers of RCTs** | Godin et al. (2011) [21], Nimavat et al. (2020) [20] | These resources provide a short checklist of simple questions addressing methodology, reporting, and interpretation for clinicians to keep in mind when appraising RCTs. | Ferreira et al. (2020) [16], Park et al. (2020) [17] |

Our proposed methodological review has two main objectives: (1) to describe ACTs in medicine and surgery and evaluate them using the CONSORT ACE guidelines, so that we can brainstorm novel risk of bias items and (2) to search and identify any current risk of bias items pertaining to ACTs. We will develop our risk-of-bias tool for ACTs in accordance with the framework described by Whiting et al. (2017) [27].

## METHODS

### Study design

We present a protocol to describe the rationale for performing a methodological review of ACTs and generate a list of bias items related to ACTs. We will follow the methodological framework proposed by Whiting et al. (2017) and Sanderson et al. (2007) [27, 28]. This protocol was written with guidance from a methodology review protocol published by Lunny and colleagues, who set out to create a novel risk-of-bias tool for network meta-analyses [29]. As described by Lunny and colleagues, subsequent steps in creating a risk-of-bias tool will include (1) a multi-round Delphi survey and panel to select, refine, and compile bias items into a single tool; (2) a pilot test to further refine the draft tool; and (3) a knowledge translation strategy to disseminate the final risk-of-bias tool [29]. These steps will be addressed in future studies as we progress through this framework in developing this proposed ACT risk-of-bias instrument. We did perform this systematic review following the PRISMA-P checklist [30].

### Eligibility

There will be three types of studies included in this scoping review. *Study type 1* will be ACTs published in the medical and/or surgical literature. A review of all ACTs investigating medical and surgical interventions will facilitate a comprehensive description of the type and quality of ACTs. With regards to the review of all published ACTs, we will perform our search to take place after that of Purjak et al. (2022) who performed a scoping review of all RCTs with adaptive designs and evaluated reporting quality [31]. *Study type 2* will be studies that describe items related to bias, reporting, or methodological quality of ACTs in any discipline. We will retain all items related to methodological bias and/or reporting as they may be able to be translated into a risk-of-bias tool. *Study type 3* will be studies that assess the methodological quality, or risk of bias, of ACTs using criteria that focus on methodological features specific to ACTs. Study type 1 articles will be categorized and analyzed separately, while study types 2 and 3 will be analyzed with the goal of collating bias items. All relevant inclusion and exclusion criteria are listed in Table 2.

**Table 2.** Study inclusion and exclusion criteria.

| Study type 1: Adaptive clinical trials (ACTs) |  |
| --- | --- |
| Inclusion criteria | Exclusion criteria |
| Any publication type (e.g., primary study, published abstract, thesis dissertation, pre-print manuscript)<br>Published in any language.<br>Published in any medical or surgical discipline<br>Evaluating at least two medical or surgical interventions<br>Randomized-controlled prospective design<br>Incorporating at least one adaptive component (e.g., adaptive randomization, Bayesian adaptive method, continued reassessment, dose response model, frequentist adaptive method, multi-arm, multi-stage [MAMS], sample size re-estimation) that is defined within the title, abstract, and/or main text of the study. | No defined adaptive design component within the title, abstract, and/or main text of the trial<br>Bayesian/adaptive subgroup analysis or re-analysis of previously collected RCT data<br>Bayesian/adaptive simulation studies with no randomized prospective patient data collection |
| Study type 2: Studies that describe items relating to methodological bias, reporting, or quality in ACTs |  |
| Inclusion criteria | Exclusion criteria |
| Any publication type (e.g., primary study, published abstract, thesis dissertation, pre-print manuscript)<br>Published in any language.<br>Any study design (i.e., RCT, prospective/retrospective cohort, cross-sectional, case series, case report, meta-analysis, scoping review, systematic review, narrative review, author editorial).<br>Mention, describe, evaluate at least one methodological bias, reporting, or quality item or tool pertaining to ACTs | Reported methodological item or tool not mentioned or described in the context of adaptive trial designs. |
| **Study type 3:** Studies that assess the methodological quality or risk of bias of ACTs |  |
| **Inclusion criteria** | **Exclusion criteria** |
| Any publication type (e.g., primary study, published abstract, thesis dissertation, pre-print manuscript)<br>Published in any language.<br>Any study design (i.e., RCT, prospective/retrospective cohort, cross-sectional, case series, case report, meta-analysis, scoping review, systematic review, narrative review, author editorial).<br>Evaluate or describe the methodological quality or risk of bias of at least one ACT | Reported methodological item or tool not mentioned or described in the context of adaptive trial designs. |

We will include all articles with any publication status and written in any language. In cases where the co-authors are not fluent or review authors are unable to understand the study text, we will utilize Google Translate (Mountain View, CA, USA). If we identify a systematic review or meta-analysis of ACTs, we will not include the systematic review in the final ACT synthesis, but instead any primary ACTs that meet inclusion criteria.

### Search Strategy

All databases to be used in this review were selected with guidance from Lunny et al. (2021) [29]. We will search all databases with no language or publication type limits. We will search the following databases: MEDLINE (Ovid), CINAHL, EMBASE (Ovid), the Cochrane library, the Cochrane Central Register of Controlled Trials (CENTRAL), Web of Science, BIOSIS, Derwent Innovations Database, and KCI. We will also search clinical trials registries including clinicaltrials.gov and the WHO International Clinical Trials Registry Platform (ICTRP). All searches for peer-review ACTs will be performed from the end date of the scoping review of ACTs published by Purja and colleagues [31]. We will search the following grey literature databases and resources: the EQUATOR network, dissertation abstracts, websites of evidence synthesis organizations (e.g., Campbell Collaboration, Cochrane Multiple Treatments Group, CADTH, NICE-DSU, Health Technology Assessment International (HTAi), Pharmaceutical Benefits Advisory Committee, Institut für Qualität und Wirtschaftlichkeit im Gesundheitswesen, European Network for Health Technology Assessment, Guidelines International Network, ISPOR, International Network of Agencies for Health Technology Assessment, and JBI), and methods collections (e.g., Cochrane Methodology Register, AHRQ Effective Healthcare Programme). We will also search LIGHTS and LATITUDES (https://www.latitudes-network.org/), which are two methodological registries that capture guidance and validity assessment tools, respectively [32]. All online registries will be searched using the following terms “adaptive clinical trial”, “bias”, and/or risk-of-bias (ROB)”. The words found within the titles, abstracts, and MeSH terms of relevant articles were used to develop focused search strategies for each database. Reference lists of studies found will also be searched for additional papers to be included. The MEDLINE search will be validated for 10 studies identified by senior authors prior to screening. Eligibility screening will only begin after these 10 trials are identified from the search strategy. All database search strategies are described in Appendix 2.

The search strategy will be generated by two authors (P.S. and D.O.) alongside a librarian specialist. It will be generated and reviewed in accordance with PRESS (Peer Review Electronic Search Strategies) guidelines [33]. Any concerns with search strategy generation will be raised with a senior methodologist (M.B.). The database search will be conducted without limitations to publication type, status, language, or date to identify existing tools or articles.

### Screening and data extraction

First, we will pilot eligibility criteria in Microsoft Excel (Redmond, WA, USA) by evaluating a sample of 25 citations amongst two independent reviewers. If high agreement is achieved (≥70%), then we will continue to abstract screening with two reviewers. If less than high agreement is achieved, then the eligibility criteria will be re-examined, and additional teaching sessions will be provided to reviewers. All screening and full-text review will be conducted using the web-based application, Covidence (http://www.covidence.org; Melbourne, Australia). Study titles and abstracts will then be assessed for relevance and eligibility. Any disagreements identified during these screening and review stages will be resolved via discussion until consensus is reached. A third senior reviewer will arbitrate if screening or full-text review disagreements cannot be resolved.

A data extraction form will be generated using Microsoft Excel and piloted by reviewers on five included studies. Two authors will extract data from all included studies. We will first categorize all sources based on our eligibility criteria and we will extract author details, publication year, and study type. For all ACTs, we will extract the following information: type of adaptive design, patient details (e.g., number of enrolled/analyzed patients, eligibility criteria, patient demographics), pre-specified interim analyses and adaptive decisions, sample size calculations, treatment group details (e.g., randomization plan, allocation sequence generation, number of treatment groups, blinding details and maintenance), outcome details (e.g., primary and secondary outcome measures, statistical analysis plan), results (e.g., baseline patient data, reported outcomes, final statistical analysis, patients lost to follow-up), and any methodological limitations identified by the authors. All risk of bias analyses will be performed by two independent reviewers using the Cochrane ROB 2.0 tool. We will also perform an assessment of reporting quality using the CONSORT ACE guidelines. All disagreements will be resolved via discussion and if consensus is not reached, then a senior reviewer (P.S.) will arbitrate the final decision. From this process of risk of bias and reporting quality assessment of published ACTs, we will brainstorm novel risk of bias items and fields.

For all studies that identified bias items, tools, or quantified methodological bias in ACTs, we will generate the following list of headings: type of tool (e.g., tool, scale, checklist, or domain-based tool); scope of the tool; number of items within the tool; domains within the tool; whether the item relates to reporting or methodological quality; ratings of items and domains within the tool; methods used to develop the tool and the availability of an “explanation and elaboration”. These fields were all derived from Lunny et al. (2021) and Page et al. (2018) [29, 34]. Data will be extracted on items that are relevant to ACTs and all items will initially be extracted verbatim.

### Data analysis and reporting

We will plan to split the analysis based upon study design: (1) ACTs will be collated and described based upon all extracted fields and (2) all studies evaluating methodological quality, and/or proposing bias tools or items will be collated and items will be grouped accordingly. Firstly, we will divide ACTs based upon their respective discipline and year of publication. We will perform descriptive analyses (i.e., proportions, mean, ranges, standard deviations), where applicable, for all extracted data fields.

Next, we will categorize all studies of methodological quality and risk-of-bias tools/items in ACTs. These studies will again undergo descriptive analyses based upon previously extracted fields. All bias items will all be mapped to corresponding domains within the CONSORT-ACE guidelines, as this is the only known quality tool specific to ACT features. If no risk-of-bias tools or relevant items are identified in our review, then we will plan to hypothesize items and formulate candidate tool. We will develop a candidate tool that will undergo refinement by the authors, and in subsequent steps will be further evaluated and refined using a Delphi consensus method prior to undergoing validation and testing. All statistical analyses will be performed using R software (version 4.3.2).

### Public and public involvement

Patient or the public were not involved in the design of this research protocol.

## DISCUSSION

A comprehensive risk-of-bias tool is needed to facilitate the publication of high-quality ACTs and any future pooled analyses of ACTs. A recent scoping review of ACTs has demonstrated poor reporting based on the CONSORT-ACE guidelines [31]. We are, therefore, going to use the framework developed by Whiting and colleagues to develop a new risk-of-bias tool for randomized trials with adaptive design features [27]. The framework developed by Whiting and colleagues has been used to develop the QUADAS tool for diagnostic test studies and a risk-of-bias tool for network meta-analyses. As AI networks and computational technologies continue to be optimized, their role in generating adaptive trial paradigms and performing statistical simulations may revolutionize the future of trial design and medical innovation [35]. We, must, therefore ensure that methodological tools are developed at a similar pace so that novel trial designs and adaptive paradigms are standardized, transparent, reproducible, and interpretable for the future.

## Data Availability

No datasets were generated or analysed during the current study. All relevant data from this study will be made available upon study completion.

## APPENDICES

**Appendix 1**. Database search strategies

## Contributors

PS, TM, HZ, MA, MKG, MB, and JP conceived the research question. PS, CL, DO, EO, TM, AT, and KN composed the manuscript. All authors edited and helped to draft the manuscript. The final version of this manuscript was approved by all authors. All authors have agreed to be held accountable for this work.

## Patient consent for publication

None

## Acknowledgements

We thank Dr. Edward Mills for his helpful manuscript comments and suggestions.

## REFERENCES

1. Masic I, Miokovic M, Muhamedagic B. Evidence based medicine - new approaches and challenges. Acta Inform Med. 2008;16(4):219–25. doi: 10.5455/aim.2008.16.219-225. PubMed PMID: 24109156; PubMed Central PMCID: PMCPMC3789163.

2. Sibbald B, Roland M. Understanding controlled trials. Why are randomised controlled trials important? BMJ. 1998;316(7126):201. doi: 10.1136/bmj.316.7126.201. PubMed PMID: 9468688; PubMed Central PMCID: PMCPMC2665449.

3. Nichol AD, Bailey M, Cooper DJ, Polar, Investigators EPO. Challenging issues in randomised controlled trials. Injury. 2010;41 Suppl 1:S20–3. Epub 20100422. doi: 10.1016/j.injury.2010.03.033. PubMed PMID: 20413119.

4. Pronk AJM, Roelofs A, Flum DR, Bonjer HJ, Abu Hilal M, Dijkgraaf MGW, et al. Two decades of surgical randomized controlled trials: worldwide trends in volume and methodological quality. Br J Surg. 2023;110(10):1300–8. doi: 10.1093/bjs/znad160. PubMed PMID: 37379487; PubMed Central PMCID: PMCPMC10480038.

5. Chapman SJ, Shelton B, Mahmood H, Fitzgerald JE, Harrison EM, Bhangu A. Discontinuation and non-publication of surgical randomised controlled trials: observational study. BMJ. 2014;349:g6870. Epub 20141209. doi: 10.1136/bmj.g6870. PubMed PMID: 25491195; PubMed Central PMCID: PMCPMC4260649.

6. Pallmann P, Bedding AW, Choodari-Oskooei B, Dimairo M, Flight L, Hampson LV, et al. Adaptive designs in clinical trials: why use them, and how to run and report them. BMC Med. 2018;16(1):29. Epub 20180228. doi: 10.1186/s12916-018-1017-7. PubMed PMID: 29490655; PubMed Central PMCID: PMCPMC5830330.

7. Pushpakom SP, Taylor C, Kolamunnage-Dona R, Spowart C, Vora J, Garcia-Finana M, et al. Telmisartan and Insulin Resistance in HIV (TAILoR): protocol for a dose-ranging phase II randomised open-labelled trial of telmisartan as a strategy for the reduction of insulin resistance in HIV-positive individuals on combination antiretroviral therapy. BMJ Open. 2015;5(10):e009566. Epub 20151015. doi: 10.1136/bmjopen-2015-009566. PubMed PMID: 26474943; PubMed Central PMCID: PMCPMC4611177.

8. Stallard N, Hampson L, Benda N, Brannath W, Burnett T, Friede T, et al. Efficient Adaptive Designs for Clinical Trials of Interventions for COVID-19. Stat Biopharm Res. 2020;12(4):483–97. Epub 20200729. doi: 10.1080/19466315.2020.1790415. PubMed PMID: 34191981; PubMed Central PMCID: PMCPMC8011600.

9. Alemayehu C, Mitchell G, Nikles J. Barriers for conducting clinical trials in developing countriesa systematic review. Int J Equity Health. 2018;17(1):37. Epub 20180322. doi: 10.1186/s12939-018-0748-6. PubMed PMID: 29566721; PubMed Central PMCID: PMCPMC5863824.

10. Madani Kia T, Marshall JC, Murthy S. Stakeholder perspectives on adaptive clinical trials: a scoping review. Trials. 2020;21(1):539. Epub 20200617. doi: 10.1186/s13063-020-04466-0. PubMed PMID: 32552852; PubMed Central PMCID: PMCPMC7301522.

11. Dimairo M, Pallmann P, Wason J, Todd S, Jaki T, Julious SA, et al. The adaptive designs CONSORT extension (ACE) statement: a checklist with explanation and elaboration guideline for reporting randomised trials that use an adaptive design. Trials. 2020;21(1):528. Epub 20200617. doi: 10.1186/s13063-020-04334-x. PubMed PMID: 32546273; PubMed Central PMCID: PMCPMC7298968.

12. Sterne JAC, Savovic J, Page MJ, Elbers RG, Blencowe NS, Boutron I, et al. RoB 2: a revised tool for assessing risk of bias in randomised trials. BMJ. 2019;366:l4898. Epub 20190828. doi: 10.1136/bmj.l4898. PubMed PMID: 31462531.

13. Jadad AR, Moore RA, Carroll D, Jenkinson C, Reynolds DJ, Gavaghan DJ, McQuay HJ. Assessing the quality of reports of randomized clinical trials: is blinding necessary? Control Clin Trials. 1996;17(1):1–12. doi: 10.1016/0197-2456(95)00134-4. PubMed PMID: 8721797.

14. Haile ZT. Critical Appraisal Tools and Reporting Guidelines. J Hum Lact. 2022;38(1):21–7. Epub 20211118. doi: 10.1177/08903344211058374. PubMed PMID: 34791933.

15. Mol BW, Lai S, Rahim A, Bordewijk EM, Wang R, van Eekelen R, et al. Checklist to assess Trustworthiness in RAndomised Controlled Trials (TRACT checklist): concept proposal and pilot. Res Integr Peer Rev. 2023;8(1):6. Epub 20230620. doi: 10.1186/s41073-023-00130-8. PubMed PMID: 37337220; PubMed Central PMCID: PMCPMC10280869.

16. Ferreira D, Barthoulot M, Pottecher J, Torp KD, Diemunsch P, Meyer N. A consensus checklist to help clinicians interpret clinical trial results analysed by Bayesian methods. Br J Anaesth. 2020;125(2):208–15. Epub 20200620. doi: 10.1016/j.bja.2020.04.093. PubMed PMID: 32571570.

17. Park JJH, Harari O, Dron L, Lester RT, Thorlund K, Mills EJ. An overview of platform trials with a checklist for clinical readers. J Clin Epidemiol. 2020;125:1–8. Epub 20200513. doi: 10.1016/j.jclinepi.2020.04.025. PubMed PMID: 32416336.

18. Jung A, Balzer J, Braun T, Luedtke K. Identification of tools used to assess the external validity of randomized controlled trials in reviews: a systematic review of measurement properties. BMC Med Res Methodol. 2022;22(1):100. Epub 20220406. doi: 10.1186/s12874-022-01561-5. PubMed PMID: 35387582; PubMed Central PMCID: PMCPMC8985274.

19. Schulz KF, Altman DG, Moher D, Group C. CONSORT 2010 statement: updated guidelines for reporting parallel group randomized trials. Ann Intern Med. 2010;152(11):726–32. Epub 20100324. doi: 10.7326/0003-4819-152-11-201006010-00232. PubMed PMID: 20335313.

20. Nimavat BD, Zirpe KG, Gurav SK. Critical Analysis of a Randomized Controlled Trial. Indian J Crit Care Med. 2020;24(Suppl 4):S215–S22. doi: 10.5005/jp-journals-10071-23638. PubMed PMID: 33354045; PubMed Central PMCID: PMCPMC7724939.

21. Godin K, Dhillon M, Bhandari M. The three-minute appraisal of a randomized trial. Indian J Orthop. 2011;45(3):194–6. doi: 10.4103/0019-5413.80036. PubMed PMID: 21559097; PubMed Central PMCID: PMCPMC3087219.

22. (2023) CASP. Randomized-controlled trial CASP tool [online] [January 12, 2024]. Available from: https://casp-uk.net/.

23. Munn Z, Barker TH, Moola S, Tufanaru C, Stern C, McArthur A, et al. Methodological quality of case series studies: an introduction to the JBI critical appraisal tool. JBI Evid Synth. 2020;18(10):2127–33. doi: 10.11124/JBISRIR-D-19-00099. PubMed PMID: 33038125.

24. (SIGN) SIGN. Randomized-controlled trial tool [online] [January 12, 2024]. Available from: https://www.sign.ac.uk/.

25. Clark E, Burkett K, Stanko-Lopp D. Let Evidence Guide Every New Decision (LEGEND): an evidence evaluation system for point-of-care clinicians and guideline development teams. J Eval Clin Pract. 2009;15(6):1054–60. doi: 10.1111/j.1365-2753.2009.01314.x. PubMed PMID: 20367705.

26. Buyse M. Limitations of adaptive clinical trials. Am Soc Clin Oncol Educ Book. 2012:133–7. doi: 10.14694/EdBook_AM.2012.32.13. PubMed PMID: 24451722.

27. Whiting P, Wolff R, Mallett S, Simera I, Savovic J. A proposed framework for developing quality assessment tools. Syst Rev. 2017;6(1):204. Epub 20171017. doi: 10.1186/s13643-017-0604-6. PubMed PMID: 29041953; PubMed Central PMCID: PMCPMC5646161.

28. Sanderson S, Tatt ID, Higgins JP. Tools for assessing quality and susceptibility to bias in observational studies in epidemiology: a systematic review and annotated bibliography. Int J Epidemiol. 2007;36(3):666–76. Epub 20070430. doi: 10.1093/ije/dym018. PubMed PMID: 17470488.

29. Lunny C, Tricco AC, Veroniki AA, Dias S, Hutton B, Salanti G, et al. Methodological review to develop a list of bias items used to assess reviews incorporating network meta-analysis: protocol and rationale. BMJ Open. 2021;11(6):e045987. Epub 20210624. doi: 10.1136/bmjopen-2020-045987. PubMed PMID: 34168027; PubMed Central PMCID: PMCPMC8231030.

30. Shamseer L, Moher D, Clarke M, Ghersi D, Liberati A, Petticrew M, et al. Preferred reporting items for systematic review and meta-analysis protocols (PRISMA-P) 2015: elaboration and explanation. BMJ. 2015;350:g7647. Epub 20150102. doi: 10.1136/bmj.g7647. PubMed PMID: 25555855.

31. Purja S, Park S, Oh S, Kim M, Kim E. Reporting quality was suboptimal in a systematic review of randomized controlled trials with adaptive designs. J Clin Epidemiol. 2023;154:85–96. Epub 20221215. doi: 10.1016/j.jclinepi.2022.12.010. PubMed PMID: 36528234.

32. Hirt J, Schonenberger CM, Ewald H, Lawson DO, Papola D, Rohner R, et al. Introducing the Library of Guidance for Health Scientists (LIGHTS): A Living Database for Methods Guidance. JAMA Netw Open. 2023;6(2):e2253198. Epub 20230201. doi: 10.1001/jamanetworkopen.2022.53198. PubMed PMID: 36787138.

33. McGowan J, Sampson M, Salzwedel DM, Cogo E, Foerster V, Lefebvre C. PRESS Peer Review of Electronic Search Strategies: 2015 Guideline Statement. J Clin Epidemiol. 2016;75:40–6. Epub 20160319. doi: 10.1016/j.jclinepi.2016.01.021. PubMed PMID: 27005575.

34. Page MJ, McKenzie JE, Higgins JPT. Tools for assessing risk of reporting biases in studies and syntheses of studies: a systematic review. BMJ Open. 2018;8(3):e019703. Epub 20180314. doi: 10.1136/bmjopen-2017-019703. PubMed PMID: 29540417; PubMed Central PMCID: PMCPMC5857645.

35. Cascini F, Beccia F, Causio FA, Melnyk A, Zaino A, Ricciardi W. Scoping review of the current landscape of AI-based applications in clinical trials. Front Public Health. 2022;10:949377. Epub 20220812. doi: 10.3389/fpubh.2022.949377. PubMed PMID: 36033816; PubMed Central PMCID: PMCPMC9414344.

